# Modification of temperature-morbidity associations by social determinants of health

**DOI:** 10.64898/2026.03.20.26348922

**Authors:** Noah Scovronick, Danlu Zhang, Zachary H. McCann, Rohan D’Souza, Morgan Lane, Rebecca Zhang, Rebecca Philipsborn, Stefanie Ebelt, Howard H. Chang

## Abstract

Exposure to high ambient temperature is responsible for more than 11,000 deaths and over 230,000 disability-adjusted life-years in the United States each year. However, which individuals and populations are most at risk, and why, is still not well understood. In 2015, a subset of “Z” diagnosis codes (or “Z-codes”) were introduced as a standardized option for healthcare providers to document the social needs and conditions of their patients. To assess heat-related risk across social determinants of health (SDoH), we leverage these codes using a dataset of patient-level emergency department (ED) visits from seven US states. Using a bi-directional, time-stratified, case-crossover design and conditional logistic regression, we compared hospital encounters for seven different health outcomes with SDoH Z-codes at discharge to a reference group matched on age, sex, race, ethnicity, year and hospital. We investigated the following Z-code domains: inadequate housing (Z59.0, Z59.1), poverty-related (Z56.0, Z59.5-Z59.7), living alone (Z60.2), institutional living (Z65.1, Z59.3), and other problems with the social environment (other Z60 sub-codes). We calculated cumulative odds ratios (ORs) for a 3-day lag change in temperature from the 95th to 50th percentile, using ZIP code-specific temperature percentiles. Among 60,557,958 ED visits with available demographic and meteorological data, 461,468 (0.8%) included a SDoH Z-code. Across temperature metrics and outcomes, patients with SDoH Z-codes consistently showed higher associations with heat than the matched reference group without SDoH Z-codes. The largest difference was for acute kidney injury, with a ratio of ORs of 1.21 (1.10,1.33) for daily mean temperature. Notable subgroup findings included elevated kidney-related risks in patients with inadequate housing or poverty-related SDoH, increased mental health risks among those living alone, and elevated cardiovascular risks in people with other problems related to the social environment.

## INTRODUCTION

Exposure to high ambient temperature is responsible for more than 11,000 deaths and over 230,000 disability-adjusted life-years in the United States each year (1). As the world continues to warm, heat exposure will intensify, increasing concerns about heat-related health risks (2); anthropogenic climate change is already responsible for hundreds of heat-related deaths in the United States every summer, and the same is true for locations around the world (3). In the United States, nine of the 10 hottest years on record have occurred since 1998 (4).

A large body of research has examined associations between adverse health events and ambient temperature (5-13). Most studies have relied on administratively collected data, which generally limits analyses of differential risk to routinely available demographic information (e.g. age, sex, race) or area-level socioeconomic indicators. As a result, these population-based studies have struggled to pinpoint which individuals and populations are most vulnerable to heat, as well as the specific factors driving that vulnerability. In particular, the link between the social determinants of health (SDoH) – the social and environmental conditions in which people live, work, learn and grow (14, 15) – and temperature-related morbidity remains poorly characterized. Much of what is known about how individual social factors other than demographics shape heat risk relies on a relatively small number of studies that collected primary data after short periods of unusually extreme heat (e.g., Chicago 1995) (16, 17), which may not be generalizable.

Recognizing the growing evidence that SDoH play a critical role in many health outcomes (14, 18-20) and in health disparities (15, 20), the healthcare system has begun to integrate social context into clinical data. A major step was the inclusion of Z-codes for SDoH in the Clinical Modification of the 10^th^ revision of the International Classification of Disease (ICD-10-CM) (21-23). ICD coding is the standardized system used by healthcare providers and administrators to record diagnoses on a patient’s medical record. Introduced in late 2015, SDoH Z-codes (Z55-Z65) provide a uniform mechanism across states and facilities to collect information on patients’ SDoH (22). Importantly, any member of a patient’s care team can record a Z-code (24), enabling broader capture of social risk information.

Although current coding practices likely underestimate SDoH, it is estimated that more than a million emergency department (ED) visit records across the US each year include at least one SDoH Z-code (21, 22, 25, 26), creating new opportunities for large-scale epidemiological research. Leveraging these data is essential for understanding how social and environmental factors interact to shape health risks. Heat is a particularly salient application, as it disproportionately affects already-disadvantaged populations (27), and because climate change is expected to exacerbate heat-related health inequities in the absence of intervention (28, 29). By identifying the SDoH that contribute to adverse heat-related health outcomes, policymakers can target the subgroups most at risk. In addition, by characterizing the value of SDoH-related risk documentation at an individual level for prevention of heat-related illness, policymakers can encourage processes and procedures that increase utilization of SDoH Z codes.

Here we compiled a dataset of patient-level ED visits from seven states (Arizona, California, Georgia, Maryland, New York, Oregon, and Utah) to investigate heat-related risk across different SDoH, as documented by Z-codes. Meteorological variables were linked to the ED dataset using patient residential ZIP codes and date of admission. For all visits and visits for specific diagnoses [respiratory diseases (RESP), cardiovascular diseases (CVD), fluid-and-electrolyte imbalance (FEI), acute kidney injury (AKI), and mental health (MENTAL)], we assessed associations with temperature using a case-crossover study design, and compared resulting odds ratios from patient visits with select Z-codes to those from a matched reference group. To our knowledge, this is the first study assessing the impacts of SDoH on ambient temperature health risks utilizing Z-codes.

## RESULTS

Across the seven states, there were 60,557,958 million warm-season ED visits during 2016-2019 with available demographic and meteorological data, of which 461,468 (0.8%) visits included a Z-code for an adverse social condition (Table 1). Patients with a SDoH Z-code were more likely to be male and White than patients without a SDoH Z-code. By far the most recorded group of SDoH Z-codes in our analysis were problems related to housing (Table 1). More detailed descriptions of the included SDoH Z-codes are available in Supplementary Table 1, and a summary of ED visits by state are reported in Supplementary Table 2.

**Table 1.** Summary of warm-season emergency department visits during 2016-2019 included in the seven-state dataset, for all visits and by *a priori* selected social determinants of health as recorded through Z-codes on the encounter record.

| ICD-10 Diagnosis Code | Short Description | Number of visits | Percent male | Percent White |
| --- | --- | --- | --- | --- |
| Any code | All visits<sup>‡</sup> | 60,557,958 | 47.1% | 49.5% |
| Z55-Z65 | Any SDoH | 461,468 | 62.2% | 50.5% |
| Z59.0, Z59.1 | Housing-related | 294,280 | 71.1% | 50.6% |
| Z56.0, Z59.5-Z59.7 | Poverty-related | 63,140 | 60.7% | 42.9% |
| Z60.2 | Living alone | 23,912 | 39.2% | 60.3% |
| Z65.1, Z59.3 | Institutional living | 9,960 | 75.9% | 36.8% |
| Other Z60 codes | Other social factors | 11,943 | 49.6% | 52.0% |

<sup>‡</sup> Within this group, 2,311,118 were selected for one of the matched reference groups.

For each outcome, higher warm-season temperatures were associated with increased ED visits in both the group of patients with a SDoH Z code and the corresponding reference group without a SDoH Z-code (matched on age, sex, race, ethnicity, year and hospital) (Figure 1). Across all six outcomes, patients with SDoH Z-codes showed higher heat risks compared to their matched reference group. The largest differences were for FEI and AKI; the ratio of odds ratios for a change in average temperature from the 50^th^ to 95^th^ temperature percentile for FEI and AKI were 1.09 (95% CI: 1.03, 1.14) and 1.21 (95% CI: 1.10, 1.33), respectively (Supplementary Table 3).

**Figure 1.**
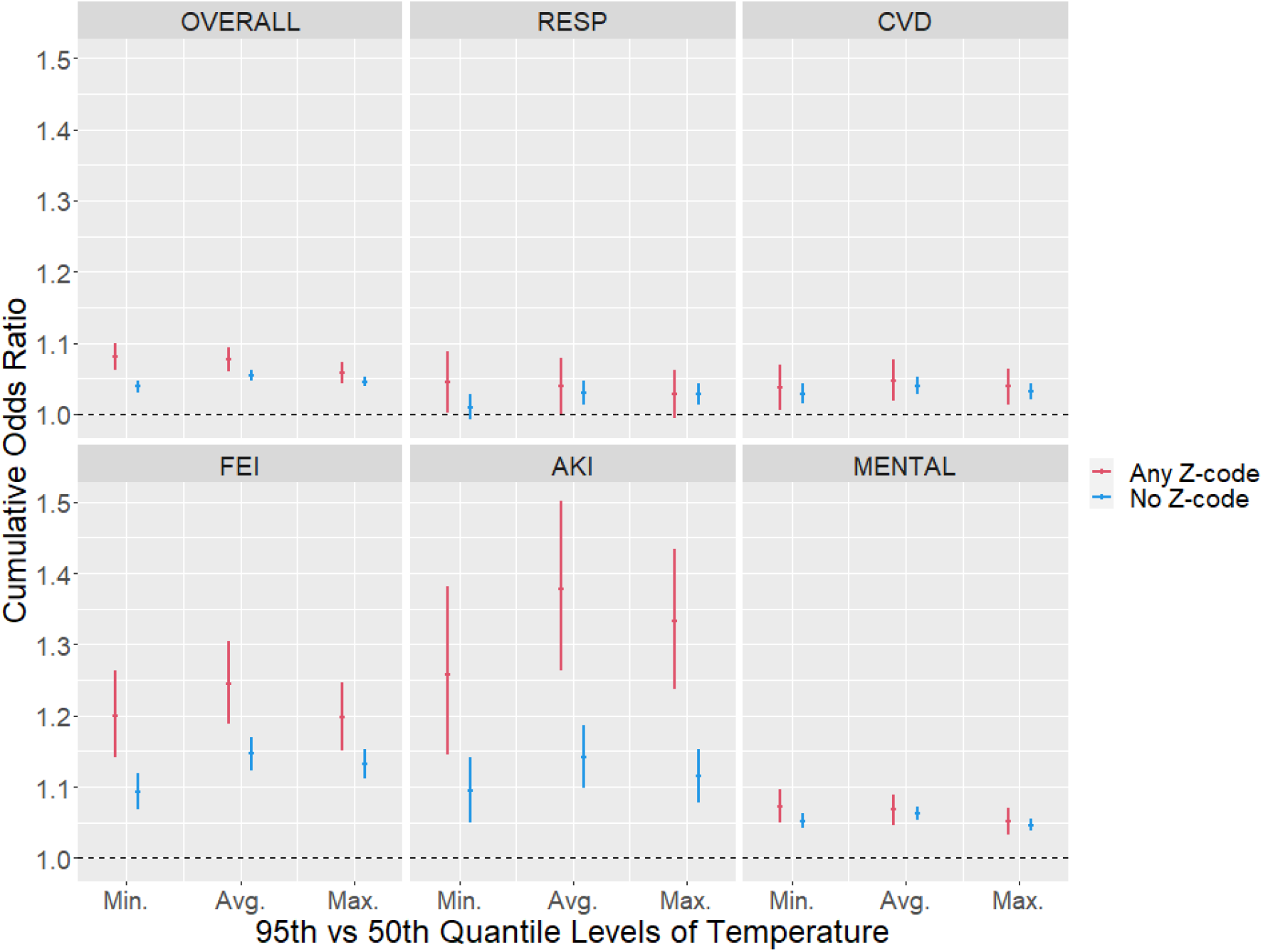
Odds ratios for three-day cumulative lag (lag 0-2) associations between warm-season emergency department visits and daily minimum, average and maximum temperature, among patient visits with any SDoH Z-code (Z55-Z65) and a matched reference group without a SDoH Z code. OVERALL = any of the specific diagnoses, RESP = respiratory, CVD = cardiovascular, FEI = fluid and electrolyte imbalance, AKI = acute kidney injury, MENTAL = mental health outcomes.

Extending the analyses to assess specific SDoH domains, patient visits with problems related to housing had high heat risks for both FEI (OR=1.31 [1.24–1.39]) and AKI (OR=1.44 [1.29–1.61]) (Figure 2), which were significantly (p<0.05) higher than their matched reference groups (also see Supplementary Table 3). For AKI, patient visits with poverty-related and social environment-related Z-codes also showed higher heat risks than their matched reference groups. Other notable results include the higher heat risk for mental health outcomes in people living alone (ratio of ORs = 1.04 [0.93-1.17]) or with other problems related to the social environment (ratio of ORs = 1.07 [0.92-1.25]), and the elevated heat risk for cardiovascular disease in those with a social environment Z-code (ratio of ORs = 1.26 [1.01-1.57]). The ratios of odds ratios for all SDoH-outcome pairs are reported in Supplementary Table 3.

**Figure 2.**
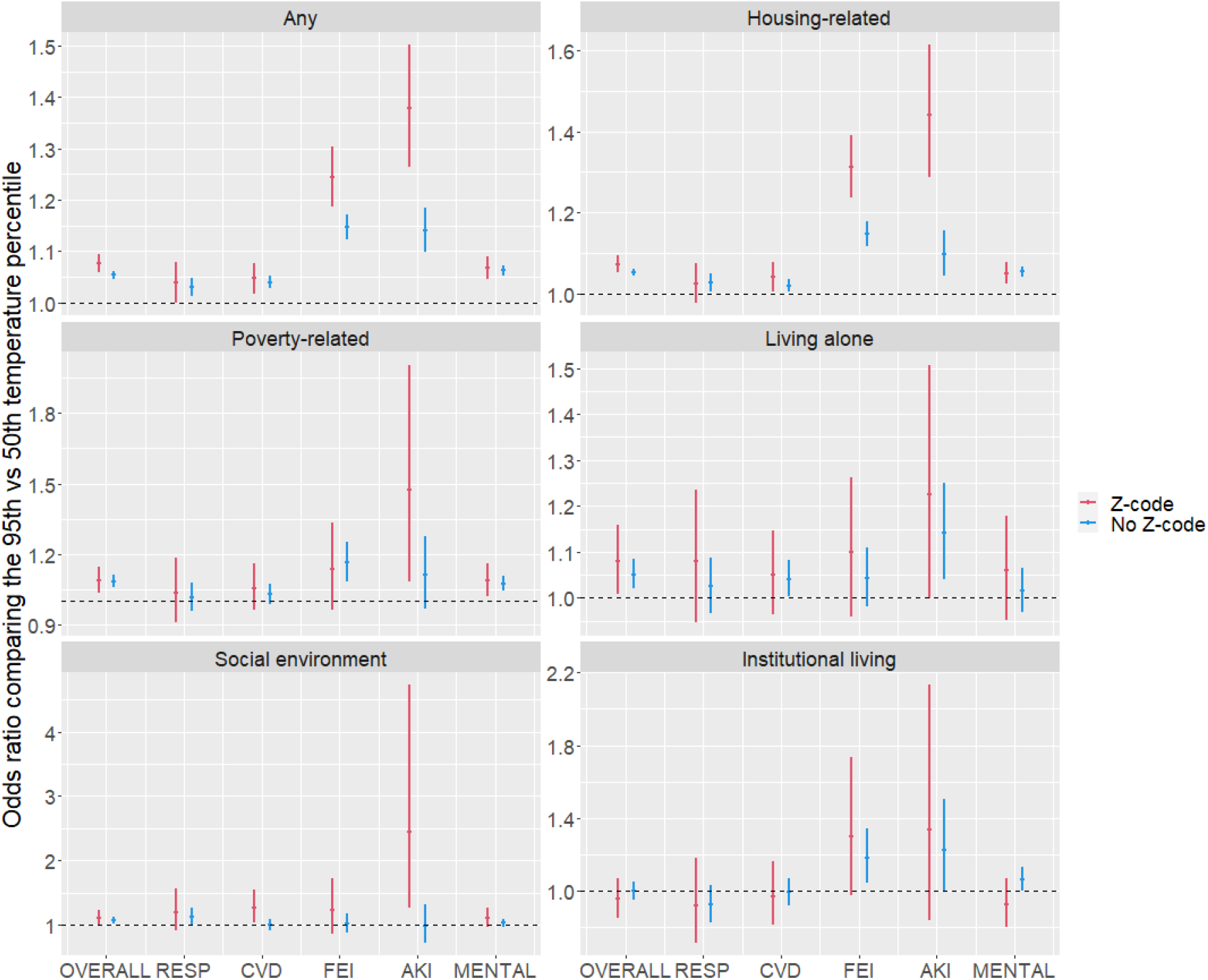
Odds ratios for three-day cumulative lag (lag 0-2) associations between warm-season emergency department visits and daily average temperature, among patient visits with select SDoH Z-codes and a matched reference group without the SDoH Z-code. OVERALL = any of the specific diagnoses, RESP = respiratory, CVD = cardiovascular, FEI = fluid and electrolyte imbalance, AKI = acute kidney injury, MENTAL = mental health outcomes.

Figure 3 displays the exposure-response curves across the distribution of average daily temperature for the main analyses comparing heat-health risks among patient visits with any SDoH Z-code to their matched reference groups. Curves were non-linear in most cases. For FEI and AKI, increases in temperature conferred a higher risk of an ED visit in both those with a SDoH Z-code and the matched reference group across much of the temperature distribution; the risk difference between the groups was most pronounced at the hottest temperatures. In contrast, for cardiovascular and mental health outcomes, the risk difference between those with a SDoH Z-code and the matched reference groups was most evident at the lower end of the warm season temperature distribution. The non-linear features can also be seen when comparing odds ratios and ratios of odds ratios at different temperature percentiles (Supplementary Table 3).

**Figure 3.**
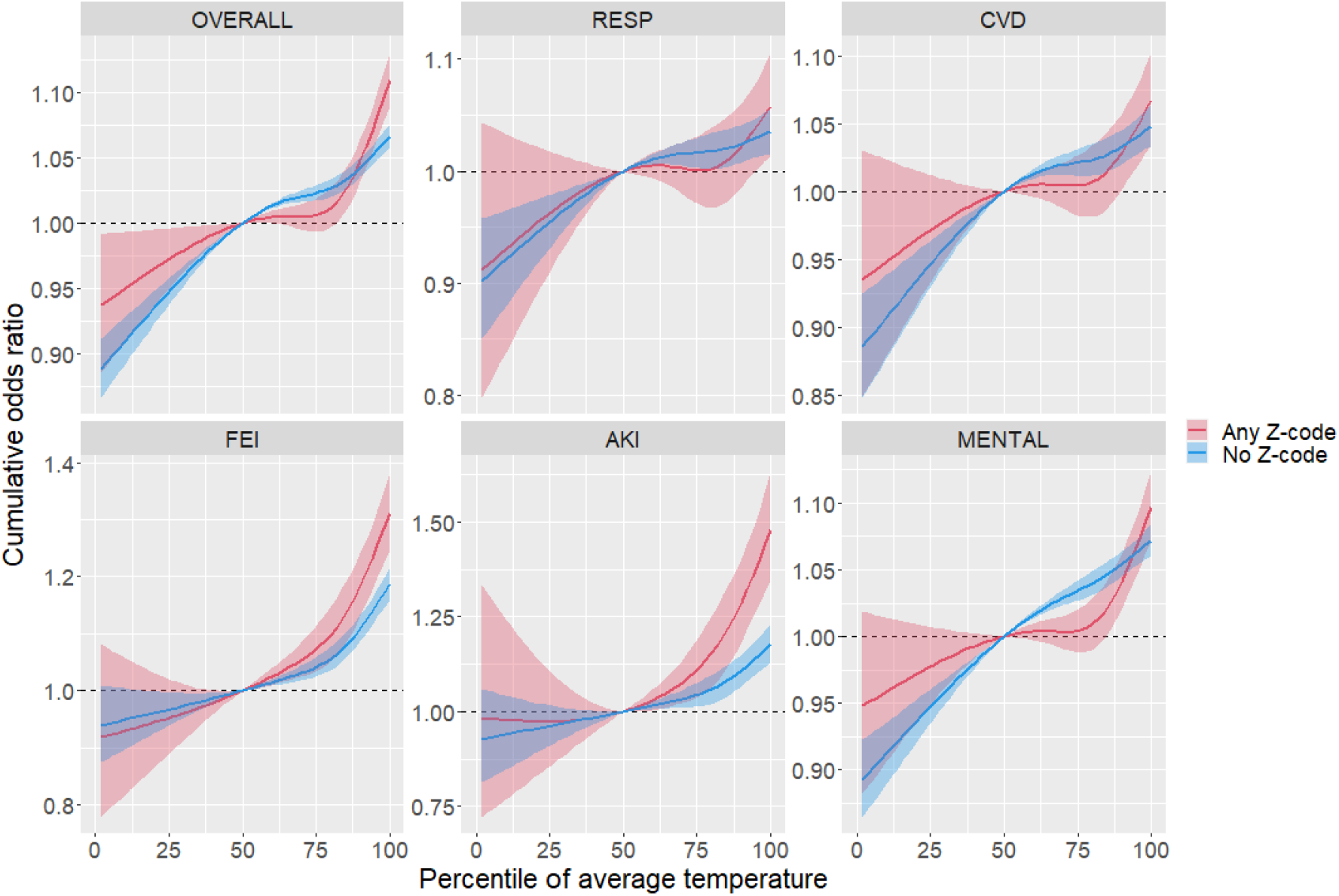
Exposure-response curves showing three-day cumulative lag (lag 0-2) associations between daily average temperature and hospital visits in patients with select Z-codes, and a matched reference group. OVERALL = any of the specific diagnoses,RESP = respiratory, CVD = cardiovascular, FEI = fluid and electrolyte imbalance, AKI = acute kidney injury, MENTAL = mental health outcomes.

## DISCUSSION

To our knowledge, this is the first study to use “Z” diagnosis codes to explore how associations between heat exposure and ED visits may be modified by the social determinants of health. We found that patients with a documented SDoH almost universally showed a higher risk of presentation to an ED in elevated heat compared to matched patients without an SDoH Z code. This was true for all five groups of specific health outcomes we evaluated. Of those five outcomes, elevated heat risks among those with a SDoH were most evident for fluid and electrolyte imbalance and acute kidney injury.

Within the different domains of SDoH, we observed variation in temperature effect sizes across health outcomes. Variation is not surprising, as the different SDoH domains represent disparate social processes. Some domains, such as experiencing homelessness or poverty may be indicative of the inability to afford material resources that may help reduce heat exposure and mitigate heat-related morbidity. Other Z-code domains, such as living alone or social isolation, may indicate that individuals do not have social networks that provide the knowledge or ability to access resources or implement adaptive strategies that might otherwise be achievable (30, 31).

The existing literature on how the SDoH affect heat-health associations is relatively small. Studies using routinely collected morbidity or mortality data have generally been limited to investigating demographic factors such as age, sex, and race, which often are not straightforward predictors of economic conditions, disease risk, and/or the need for social support. That literature finds a relatively robust link between age and heat vulnerability, particularly for mortality, but studies on race and sex are somewhat more mixed and possibly dependent on the location and health outcome (6, 12, 32-37). There are a small number of studies from the US using educational attainment as a proxy for SDoH, which suggest higher levels of education may be protective (38, 39).

There have been several studies, often case-control studies, that have explored risk factors for ill-health – mainly mortality – during periods of extreme heat. For example, living alone was associated with a higher risk of heat-related death during heat waves in Chicago in the 1990s; quality housing, such as air conditioning and certain structural characteristics, was protective (17, 40). Similar findings were reported during heatwaves in Australia and Europe (16, 41). Several of these studies also found that participating in social activities and having more social contacts is protective (17, 40, 41). All of these results are broadly consistent with our findings.

This study has several limitations. Some of the limitations are present in any study of this type, such as those associated with using temperature data derived from station measurements, and the fact that ambient temperature at a residential location is not a perfect proxy for personal exposure (42-45). In our study, exposure misclassification may occur disproportionately in Z-coded patients with issues related to housing and homelessness, as reported ZIP codes may represent places that are not where the person is living (e.g., service providers, addresses of friends or family, or a PO Box).

There are also other challenges specific to our use of Z-codes to study heat-health relationships. For example, it is widely believed that SDoH Z-codes have been under-utilized (22, 46), which leads to underreporting of SDoH. We accounted for this issue to some extent by including hospital as a matching factor. Thus, the comparative analysis, comparing results from those with and without SDoH Z-codes, was limited to facilities that recorded Z-codes during the study period. Regardless, under-reporting in our context likely biases the difference in effect sizes between the Z-coded and reference group towards the null, as some patients who should have one or more SDoH Z-code would end up in the reference group. As Z-coding becomes more engrained in hospital recordkeeping, issues related to underreporting should decrease and the power of this methodology to support decision-making will increase.

There are also limitations related to our choice of Z-code SDoH domains. We chose five domains for this first epidemiological assessment of heat risk based on the existing literature, but there are other possible domains we did not investigate, including issues related to education/literacy or drug abuse; these factors may also confer heat risk (32, 47). In some cases, we grouped codes into imperfect categories, for example the institutional living category, which includes problems related to living in a residential institution as well as imprisonment and other incarceration. The categories were designed to include different codes that may work through related pathways and where grouping would aid with statistical power. In the future, as more years of data become available, especially data after the Covid-19 years (which severely affected hospital visit rates and patient composition), it should be possible to explore individual Z codes with more statistical power. In general, future research investigating the strengths and limitations of applying Z-codes will lead to more refined epidemiological applications.

The finding that heat risk is elevated among individuals with diverse SDoH underscores the need for multiple heat intervention points and suggests that heat protection strategies must be tailored to specific populations. Determining how to prioritize and evaluate these options – ranging from measures to reduce exposure (e.g., housing improvements or urban design) to interventions that enhance social support or heat-health literacy – remains an important area for future research.

One practical implication of this study is that healthcare teams are well positioned to identify and counsel patients at risk for heat-related illness, to connect patients to available social services, and to advocate for systems and strategies to protect patients in extreme heat. As an example, SDoH documentation in the electronic medical record could flag at-risk patients to receive notifications during extreme heat with guidance on how to stay safe, leveraging SDoH Z codes for early warning systems. Recognition and documentation of SDoH by a healthcare provider could serve as an important indicator of heat risk, and this signal will likely become more actionable as systematic reporting of patient SDoH becomes more widespread.

## METHODS

### Health Data

Patient-level emergency department (ED) discharge data were obtained from seven geographically and climatically diverse states: Arizona, California, Georgia, Maryland, New York, Oregon, and Utah during some or all of 2016-2019 (Supplementary Table 4). These data include any ED visit whether the patient was admitted, or seen in the ED and discharged (i.e., not admitted).

Each record includes the visit date, a facility (hospital) identifier, patient ZIP code of residence, age or age group at the time of the visit, self-reported race and/or ethnicity, and sex. Records also report primary and secondary ICD-10 diagnosis codes, which we used to identify health outcomes of interest as well as social determinants of health through Z-codes. ED visit records typically list one primary diagnosis and multiple secondary diagnoses; Z-codes are generally recorded as secondary diagnoses and therefore do not represent the primary reason for the visit.

Outcomes of interest were selected based on prior literature on heat-health relationships (e.g., 9, 10, 13, 33, 37, 48, 49), namely respiratory diseases (RESP, ICD-10 J00-J99), cardiovascular diseases (CVD, ICD-10 I00-I99), fluid-and-electrolyte imbalance (FEI, ICD-10 E86-E87), acute kidney injury (AKI, ICD-10 N17), and mental health (MENTAL, ICD-10 F01-F99). Each outcome was identified using primary or secondary ICD-10 diagnosis codes. We also conduct analyses with all of these health outcomes together, as an aggregate group of heat-sensitive diseases (OVERALL).

Based on the existing literature – primarily from case control studies conducted during heat waves (16, 17, 40, 41) – we selected five SDoH domains for analysis, defined as follows:

- Housing-related (ICD-10 codes Z59.0, Z59.1), which includes homelessness or other inadequate housing
- Poverty-related (Z56.0, Z59.5-Z59.7), which includes problems related to poverty, low income, unemployment, and inadequate welfare support
- Living alone (Z60.2)
- Institutional living (Z65.1, Z59.3), which includes problems related to residential institutions and imprisonment
- Other problems related to the social environment (other Z60 sub-codes), which includes several problems related to acculturation and social exclusion.

Exact descriptions of each SDoH Z-code can be found in Supplementary Table 1.

### Meteorological Data

Daily minimum and maximum temperature, and daily average dew-point temperature were obtained from the Daymet product (50), which provides data at 1 km resolution. Daily mean temperature was calculated as the average of minimum and maximum temperature. The meteorological data was merged with the health data based on the centroid of each patient’s residential ZIP code. To facilitate the combining of information across locations in the statistical modeling, we converted the daily absolute temperature values to ZIP code-specific temperature percentiles using warm-season data over all study years. Average minimum, mean and maximum temperatures by state during the study period can be found in Supplementary Table 5.

The analysis was restricted to the warm season (May – October).

### Frequency Matching

Across all seven states, there were 60,557,958 warm-season ED visits with complete demographic characteristics and available meteorological data; 461,468 (0.8%) visits had a SDoH Z-code on their encounter record.

ED visit patients with and without a SDoH Z-code had different demographic distributions. To allow for an appropriate comparison of the effect of temperature on these two populations – those with the SDoH Z-codes and those without – we conducted frequency matching based on age group (<18, 18-49, 50-64, 65-74, and 75+), sex, race/ethnicity (Asian, Black, Hispanic, White and others), facility, and year of visit.

Frequency matching was done for each combination of ED visit outcome (OVERALL, RESP, CVD, FEI, AKI, and MENTAL) and specific SDoH Z-code. Since ED visits with a SDoH Z-code represented small percentage of all ED visits, a 1:5 matching was used to gain statistical power. Specifically, for each ED visit with a SDoH Z-code, we randomly selected five ED visits that had no SDoH Z-code and shared the same health outcome and other matching variables. For example, an ED visit by 66-year-old White male with a primary AKI diagnosis and a given SDoH Z-code would be matched with five patient visits made at the same hospital in the same year who had primary AKI diagnoses and were also White, male, and aged 65-74. We only included ED visits with a SDoH Z-code for whom there were five matched visits available. For matching, we treated primary and secondary diagnoses separately, matching patients only when diagnoses appeared in the same position (primary with primary or secondary with secondary), but not across positions.

Of the 60,557,958 warm-season ED visits, 35,689,261 visits were potential matches for at least one SDoH Z-coded visit. Ultimately, using the 1:5 matching ratio, 2,311,118 visits were selected as a match for use in at least one of the epidemiologic models.

### Statistical Modeling

To assess associations of temperature with ED visits, we applied a bidirectional time-stratified case-crossover design. For each case (i.e., ED visit), control days were chosen as the same day of the week within the same month and year, leading to 3-4 control days per case. This approach inherently controls for long-term trends, seasonality, and day of the week. Conditional logistic regression was used to estimate non-linear associations (odds ratios) between ED visits and temperature at up to two days of lag, as follows:

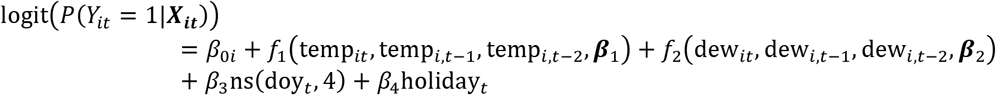

where *p*(Y_*it*=_1|X_*t*_)denotes the probability of having an ED visit on day t for patient *i*given exposures and covariates X_i*t*_. Exposures were defined as temperature percentiles on day t at the residential ZIP code of patient *i*(temp_*it*_), and 1-day and 2-day lagged temperature percentiles at the same ZIP-code (temp_*i*,t-1_and temp_*i,t*−2_). Covariates included ZIP-code level dew-point temperature on lag 0-2 days (dew_*it*_, dew_*i,t*−1_, and dew_*i,t*−*2*_). An additional within-month trend was accounted for smoothly using natural cubic splines for day of year (doy_*t*_) with 4 degrees of freedom, and an indicator of federal holidays on day t. Non-linear associations for lagged temperature and dew-point temperature (*f*_1_ and *f*_2_) were specified as natural cubic splines with 3 degrees of freedom and were additive across lag days.

We ran separate models for the group with a given SDoH Z-code and its matched reference group, and compared odds ratios and ratios of odds ratios.

We selected two exposure contrasts to summarize the estimated non-linear temperature effects: (1) the risk at the 95^th^ versus 50^th^ temperature percentiles, and (2) the risk at the 75^th^ versus 25^th^ temperature percentiles.

All analyses were performed using R 4.2.0 (51).

This study was approved by Emory University’s Institutional Review Board (Protocol 2025P009931).

## Supporting information

Supplemental Exhibits

## Data Availability

We cannot make the hospital records data (neither patient-level nor aggregate forms) available to external investigators given restrictions in our data use agreements. All temperature data is publicly available.

https://daymet.ornl.gov/

## ACKNOWLEDGEMENTS

This study was funded through two grants from the US National Institute of Environmental Health (R21ES034190, R01ES027892). We are grateful for the support of the health data sources listed in the following sentence and their contributing hospitals. The ED visits data used to produce this publication were acquired from the Arizona Department of Health Services; California Department of Health Care Access and Information; Georgia Hospital Association; Maryland Department of Health, Health Services Cost Review Commission; Oregon Healthcare Enterprises, Inc., Apprise Health Insights, a subsidiary of the Oregon Association of Hospitals & Health Systems; New York State Department of Health, Statewide Planning and Research Cooperative Systems (SPARCS); and Utah Department of Health, Office of Health Care Statistics (OHCS). The analysis, interpretation, conclusions, and views presented in this paper are solely those of the authors and do not reflect the official positions or conclusions of the listed data sources. The authorization to release this information does not imply any endorsement of the study or its findings by these data sources. The data sources, along with their employees, officers, and agents, do not make any representations, warranties, or guarantees regarding the accuracy, completeness, timeliness, or suitability of the information presented here.

