## Supplemental Exhibits for "Modification of temperature-morbidity associations by social determinants of health"

**Supplementary Table 1. Description of individual Z-codes for social determinants of health.**

| ICD Code | Description |
|---|---|
| Any code | All visits |
| Any Z55-Z65 code | Any social determinant of health code |
| **Housing-related** | |
| Z59.0 | Homelessness |
| Z59.1 | Inadequate housing, unspecified |
| **Poverty-related** | |
| Z56.0 | Problems related to employment and unemployment |
| Z59.5 | Extreme poverty |
| Z59.6 | Low income |
| Z59.7 | Insufficient social insurance and welfare support |
| **Living alone** | |
| Z60.2 | Problems related to living alone |
| **Institutional living** | |
| Z65.1 | Imprisonment and other incarceration |
| Z59.3 | Problems related to living in residential institution |
| **Other social factors** | |
| Z60.0 | Problems of adjustment to life-cycle transitions |
| Z60.3 | Acculturation difficulty |
| Z60.4 | Social exclusion and rejection |
| Z60.5 | Target of (perceived) adverse discrimination and persecution |
| Z60.8 | Other problems related to social environment |
| Z60.9 | Problem related to social environment, unspecified |

**Supplementary Table 2. Number of emergency department visits in each state in the final dataset.**

| ICD Code | Description | Arizona | California | Georgia | Maryland | New York | Oregon | Utah |
|---|---|---|---|---|---|---|---|---|
| Any code | All visits | 2967176 | 13712064 | 5961032 | 770127 | 9489668 | 1913427 | 875767 |
| Any Z55-Z65 code | Any social determinant of health | 48338 | 130595 | 60321 | 13157 | 146534 | 45581 | 16942 |
| Z59.0, Z59.1 | Homeless / inadequate housing | 35106 | 80290 | 39386 | 8140 | 83349 | 37577 | 10432 |
| Z56.0, Z59.5-Z59.7 | Poverty-related | 4307 | 8395 | 4336 | 2116 | 39614 | 1000 | 3372 |
| Z60.2 | Living alone | 1572 | 12265 | 2655 | 551 | 5482 | 993 | 394 |
| Z65.1, Z59.3 | Institutional living | 1284 | 5208 | 916 | 48 | 2145 | 228 | 131 |
| Other Z60 codes | Other social factors | 596 | 1992 | 1483 | 744 | 6362 | 570 | 196 |

**Supplementary Table 3. Odds ratios and ratios of odds ratios for three day (lag 0-2) cumulative lagged associations between emergency department visits and daily average temperature at two exposure contrasts.**

| Outcome | SDoH cateogry | OR 95$^{th}$ vs 50$^{th}$ Z-code | OR 95$^{th}$ vs 50$^{th}$ No Z-code | Ratio of ORs 95$^{th}$ vs 50$^{th}$ No Z-code | OR 75$^{th}$ vs 50$^{th}$ Z-code | OR 75$^{th}$ vs 50$^{th}$ No Z-code | Ratio of ORs 75$^{th}$ vs 50$^{th}$ No Z-code |
|---|---|---|---|---|---|---|---|
| Any of the specific diagnosis | Housing-related | 1.07 (1.05, 1.09) | 1.05 (1.04, 1.06) | 1.02 (1, 1.04) | 1.01 (0.97, 1.05) | 1.08 (1.07, 1.1) | 0.93 (0.9, 0.97) |
| | Poverty-related | 1.09 (1.04, 1.15) | 1.09 (1.06, 1.11) | 1.00 (0.95, 1.06) | 1.09 (1, 1.19) | 1.14 (1.1, 1.18) | 0.96 (0.87, 1.05) |
| | Living alone | 1.08 (1.01, 1.16) | 1.05 (1.02, 1.08) | 1.03 (0.95, 1.11) | 0.99 (0.89, 1.11) | 1.07 (1.02, 1.12) | 0.93 (0.82, 1.05) |
| | Other social environment | 1.11 (1, 1.24) | 1.07 (1.02, 1.13) | 1.03 (0.92, 1.17) | 0.91 (0.75, 1.09) | 1.04 (0.96, 1.13) | 0.87 (0.71, 1.07) |
| | Institutional living | 0.96 (0.85, 1.07) | 1.00 (0.95, 1.05) | 0.96 (0.84, 1.08) | 1.09 (0.91, 1.31) | 1.10 (1.02, 1.18) | 0.99 (0.82, 1.21) |
| | Any SDoH | 1.08 (1.06, 1.09) | 1.05 (1.05, 1.06) | 1.02 (1, 1.04) | 1.03 (1.01, 1.06) | 1.08 (1.07, 1.09) | 0.96 (0.93, 0.99) |
| Respiratory | Housing-related | 1.02 (0.98, 1.07) | 1.03 (1.01, 1.05) | 1 (0.95, 1.05) | 1.01 (0.93, 1.09) | 1.10 (1.06, 1.14) | 0.92 (0.84, 1.00) |
| | Poverty-related | 1.04 (0.91, 1.18) | 1.02 (0.96, 1.08) | 1.02 (0.88, 1.18) | 1.07 (0.87, 1.33) | 1.11 (1.01, 1.22) | 0.97 (0.77, 1.22) |
| | Living alone | 1.08 (0.95, 1.23) | 1.02 (0.97, 1.09) | 1.05 (0.91, 1.22) | 1.00 (0.81, 1.23) | 1.02 (0.93, 1.11) | 0.98 (0.78, 1.23) |
| | Other social environment | 1.2 (0.91, 1.57) | 1.13 (0.99, 1.27) | 1.06 (0.79, 1.44) | 1.14 (0.7, 1.85) | 1.17 (0.95, 1.44) | 0.98 (0.58, 1.65) |
| | Institutional living | 0.92 (0.72, 1.18) | 0.92 (0.83, 1.03) | 0.99 (0.76, 1.3) | 1.11 (0.76, 1.64) | 0.99 (0.84, 1.18) | 1.12 (0.74, 1.71) |
| | Any SDoH | 1.04 (1.00, 1.08) | 1.03 (1.01, 1.05) | 1.01 (0.97, 1.05) | 1.04 (0.98, 1.11) | 1.06 (1.04, 1.09) | 0.98 (0.91, 1.05) |

| | | | | | | | |
|---|---|---|---|---|---|---|---|
| **Cardiovascular** | Housing-related | 1.04 (1, 1.08) | 1.02 (1, 1.04) | 1.02 (0.98, 1.06) | 1.01 (0.95, 1.07) | 1.1 (1.07, 1.13) | 0.92 (0.86, 0.98) |
| | Poverty-related | 1.06 (0.96, 1.16) | 1.03 (0.99, 1.07) | 1.03 (0.93, 1.14) | 1.07 (0.91, 1.24) | 1.11 (1.04, 1.19) | 0.96 (0.81, 1.13) |
| | Living alone | 1.05 (0.96, 1.15) | 1.04 (1, 1.08) | 1.01 (0.92, 1.11) | 1.01 (0.88, 1.15) | 1.05 (0.99, 1.11) | 0.96 (0.83, 1.11) |
| | Other social environment | 1.27 (1.04, 1.55) | 1 (0.92, 1.1) | 1.26 (1.01, 1.57) | 0.99 (0.7, 1.4) | 1.04 (0.89, 1.22) | 0.96 (0.65, 1.4) |
| | Institutional living | 0.97 (0.81, 1.16) | 0.99 (0.92, 1.07) | 0.98 (0.81, 1.19) | 1.1 (0.83, 1.47) | 1.08 (0.96, 1.22) | 1.02 (0.75, 1.39) |
| | Any SDoH | 1.05 (1.02, 1.08) | 1.04 (1.03, 1.05) | 1.01 (0.98, 1.04) | 1.03 (0.99, 1.08) | 1.08 (1.06, 1.1) | 0.96 (0.91, 1.01) |
| **Fluid and Electrolyte Imbalance** | Housing-related | 1.31 (1.24, 1.39) | 1.15 (1.12, 1.18) | 1.14 (1.07, 1.22) | 1.1 (0.99, 1.23) | 1.10 (1.05, 1.15) | 1 (0.89, 1.13) |
| | Poverty-related | 1.13 (0.96, 1.34) | 1.16 (1.08, 1.25) | 0.97 (0.81, 1.16) | 1.04 (0.8, 1.34) | 1.02 (0.91, 1.14) | 1.02 (0.77, 1.35) |
| | Living alone | 1.1 (0.96, 1.26) | 1.04 (0.98, 1.11) | 1.05 (0.91, 1.23) | 1.23 (0.98, 1.55) | 0.97 (0.89, 1.07) | 1.27 (0.99, 1.62) |
| | Other social environment | 1.23 (0.87, 1.72) | 1.02 (0.88, 1.19) | 1.2 (0.83, 1.74) | 1.09 (0.6, 1.97) | 0.85 (0.66, 1.09) | 1.28 (0.67, 2.43) |
| | Institutional living | 1.3 (0.97, 1.73) | 1.18 (1.04, 1.34) | 1.1 (0.8, 1.5) | 1.06 (0.69, 1.65) | 1.14 (0.94, 1.39) | 0.93 (0.58, 1.5) |
| | Any SDoH | 1.24 (1.19, 1.3) | 1.15 (1.12, 1.17) | 1.09 (1.03, 1.14) | 1.13 (1.04, 1.22) | 1.08 (1.04, 1.11) | 1.05 (0.96, 1.14) |
| **Acute Kidney Injury** | Housing-related | 1.44 (1.29, 1.61) | 1.1 (1.04, 1.15) | 1.31 (1.16, 1.49) | 1.14 (0.93, 1.41) | 1.06 (0.97, 1.16) | 1.08 (0.86, 1.36) |
| | Poverty-related | 1.47 (1.08, 2.01) | 1.11 (0.97, 1.28) | 1.32 (0.94, 1.85) | 1.33 (0.8, 2.21) | 0.93 (0.74, 1.17) | 1.42 (0.81, 2.49) |
| | Living alone | 1.23 (1.00, 1.51) | 1.14 (1.04, 1.25) | 1.08 (0.86, 1.35) | 0.86 (0.61, 1.21) | 1.07 (0.92, 1.24) | 0.81 (0.56, 1.17) |
| | Other social environment | 2.44 (1.26, 4.72) | 0.98 (0.73, 1.33) | 2.49 (1.21, 5.15) | 1.71 (0.51, 5.69) | 0.88 (0.54, 1.46) | 1.93 (0.53, 7.11) |
| | Institutional living | 1.34 (0.84, 2.13) | 1.23 (1, 1.5) | 1.09 (0.66, 1.81) | 1.37 (0.64, 2.9) | 1.2 (0.86, 1.68) | 1.14 (0.5, 2.6) |

| | | | | | | | |
|---|---|---|---|---|---|---|---|
| | Any SDoH | 1.38 (1.26, 1.5) | 1.14 (1.10, 1.19) | 1.21 (1.10, 1.33) | 1.14 (0.98, 1.32) | 1.09 (1.02, 1.16) | 1.05 (0.89, 1.24) |
| **Mental health** | Housing-related | 1.05 (1.02, 1.08) | 1.05 (1.04, 1.07) | 1.00 (0.97, 1.02) | 1.00 (0.96, 1.04) | 1.08 (1.06, 1.1) | 0.92 (0.88, 0.97) |
| | Poverty-related | 1.09 (1.02, 1.16) | 1.07 (1.04, 1.11) | 1.01 (0.94, 1.09) | 1.09 (0.98, 1.22) | 1.08 (1.03, 1.13) | 1.01 (0.9, 1.14) |
| | Living alone | 1.06 (0.95, 1.18) | 1.02 (0.97, 1.06) | 1.04 (0.93, 1.17) | 1.04 (0.88, 1.24) | 0.99 (0.92, 1.06) | 1.06 (0.88, 1.28) |
| | Other social environment | 1.11 (0.97, 1.27) | 1.04 (0.97, 1.1) | 1.07 (0.92, 1.25) | 0.89 (0.71, 1.12) | 1.15 (1.04, 1.28) | 0.78 (0.61, 1) |
| | Institutional living | 0.93 (0.8, 1.07) | 1.06 (1.00, 1.13) | 0.87 (0.74, 1.02) | 1.02 (0.81, 1.29) | 1.11 (1.01, 1.22) | 0.92 (0.72, 1.18) |
| | Any SDoH | 1.07 (1.05, 1.09) | 1.06 (1.05, 1.07) | 1.00 (0.98, 1.03) | 1.03 (0.99, 1.06) | 1.09 (1.07, 1.11) | 0.94 (0.91, 0.98) |

**Supplementary Table 4. Time period and source of emergency department data.**

| | Years included | Data Source |
|---|---|---|
| **Arizona** | 2016-2019 | Arizona Department of Health Services |
| **California** | 2016-2019 | California Department of Health Care Access and Information |
| **Georgia** | 2016-2019 | Georgia Healthcare Association |
| **Maryland** | 2016 | Health Services Cost Review Commission |
| **New York** | 2016-2019 | Maryland Department of Health |
| **Oregon** | 2016-2019 | Statewide Planning and Research Cooperative System |
| **Utah** | 2016-2019 | New York State Department of Health |

**Supplementary Table 5. Average daily warm-season (May-October) minimum, mean and maximum temperature, by state.** The analysis was based on temperatures at the ZIP code level.

| | Average minimum temperature (°C) | Average mean temperature (°C) | Average maximum temperature (°C) |
|---|---|---|---|
| **Arizona** | 17.94 | 25.93 | 33.93 |
| **California** | 13.99 | 20.89 | 27.78 |
| **Georgia** | 18.47 | 24.49 | 30.51 |
| **Maryland** | 16.43 | 21.52 | 26.61 |
| **New York** | 12.20 | 17.76 | 23.32 |
| **Oregon** | 8.92 | 15.83 | 22.75 |
| **Utah** | 9.50 | 17.35 | 25.20 |
